# Facial video photoplethysmography for measuring average and instantaneous heart rate: a pilot validation study

**DOI:** 10.1101/2025.02.13.25322005

**Authors:** Leszek Pstras, Tymoteusz Okupnik, Beata Ponikowska, Bartlomiej Paleczny

## Abstract

**Introduction:** Video photoplethysmography (vPPG) is a contactless optical technique of recording blood pulsations in the skin vasculature using a digital camera, which is increasingly used to measure or estimate various physiological parameters. Here, we evaluate the accuracy of heart rate (HR) measurements performed using facial vPPG technology Shen.AI Vitals and a smartphone camera.

**Methods:** We studied 35 healthy volunteers in a sitting position (median age 25 years, 17 females). Video recordings of participants’ faces were obtained using the front camera of a smartphone mounted on a tripod. In parallel, a 1-lead chest electrocardiogram (ECG) was recorded to obtain reference HR values (average value from the entire 60-second measurement and multiple values averaged over 10-second or 4-second periods during the measurement).

**Results:** The mean absolute errors were 0.1, 0.2, and 0.4 beats per minute (bpm) for HR averaged over 60-second, 10-second, and 4-second periods, respectively. The errors did not exceed 1 bpm in 100.0%, 99.8%, and 94.5% of cases, respectively. Regardless of the HR averaging time, the correlation between the vPPG-based and reference values was very strong (r > 0.99, P < 0.001).

**Conclusions:** In predominantly young, white, sitting subjects, the tested vPPG technology provided highly accurate HR values, both when averaging over 60 seconds as well as in the case of short-term averages over 10 seconds or instantaneous HR values averaged over 4 seconds. The results should be confirmed in a larger study with greater diversity in age, skin tone, and lighting conditions.

## INTRODUCTION

Heart rate (HR), or pulse rate, is one of the vital signs used in medical examinations for the basic assessment of the condition of the cardiovascular system. Resting HR can help detect various diseases and predict cardiovascular and all-cause mortality [1, 2]. In particular, a UK Biobank study on over 500,000 individuals followed for up to 12 years showed that a 10-bpm increase in resting HR was associated with a 22% and 19% greater risk for all-cause mortality in men and women, respectively [3]. A meta-analysis including over 1.2m individuals followed for up to 40 years showed that higher resting HR was associated with increased cardiovascular and all-cause mortality, independent of traditional cardiovascular risk factors [4]. Moreover, an increase in resting HR over time has also been shown to be associated with higher all-cause mortality, with the risk of death increasing by 33% for every 10-bpm increase over six years [5]. Therefore, regular HR assessment is an important part of prevention [6, 7]. HR also reflects the body’s response to stress, emotions, exercise, or other stimuli, and hence it can be used for self-monitoring in both health and wellness context [8]. It can also be used for monitoring of the cardiac rehabilitation process [9, 10] as well as to monitor fatigue and recovery process in athletes to prevent overtraining and optimize training effectiveness [11–13]. Moreover, it be used to monitor drivers’ physiological state [14, 15] or to monitor human-computer interactions [16].

Photoplethysmography (PPG), i.e. an optical technique of detecting blood pulsations in the skin vasculature, is commonly used for measurements or continuous monitoring of HR and other vital signs by means of special probes/clips attached typically to a finger or ear lobe [17]. PPG sensors are also increasingly being integrated into a variety of wearable devices, such as smartwatches or wristbands [18]. The two main limitations of classic PPG-based measurements are: 1) the need for a special device, such as a pulse oximeter or a wearable equipped with PPG technology, which naturally limits the availability of such measurements, and 2) the need for skin contact, which is usually not as important in the case of self-measurements using one’s own device, but may be more important in healthcare facilities or in the case of measurements taken by another person or using someone else’s device, especially in the event of an epidemic.

The answer to the above limitations may be video-based PPG (vPPG), i.e. a remote PPG technique, also known as remote or imaging photoplethysmography, that uses digital video images of the skin to detect tiny changes in skin colour caused by blood pulsations in superficial blood vessels and the resulting changes in the blood absorption of light incident on the skin (mainly by haemoglobin) [19]. Such vPPG measurements could be performed using special light sources (e.g. LEDs with specific wavelengths) and a computer software to analyse video images from professional digital cameras.

However, such measurements can also be performed using ambient, white light (natural or artificial) as the source of light illuminating the skin, a consumer-grade camera integrated into a smartphone as the image sensor, and the smartphone processing power to analyse video images using a mobile app, thus making this technology accessible to most smartphone users without the need for any other device and without requiring skin contact. The possibility of using vPPG for contactless monitoring of HR and other vital signs has attracted a lot of attention in recent years [20–24], including the possibility of remote measurement of vital signs in telemedicine applications [25], for patient triage purposes [26, 27], or for monitoring drivers [28].

In this study, we investigated vPPG technology developed by MX Labs (Tallinn, Estonia) called Shen.AI Vitals. This technology uses face detection and tracking algorithms to obtain vPPG signals from several regions of facial skin during a 1-min video recording and then employs various signal processing algorithms to analyse and combine information from these signals (in the red, green, and blue channels) to estimate HR as well as other physiological parameters. In particular, two types of HR values are provided – after the measurement, HR averaged over the entire 1 minute is provided, whereas during the measurement (every 1 second), average values from shorter periods are provided, i.e. the average HR from the previous 10 seconds (default) or the previous 4 seconds (optional).

The aim of our study was to assess the accuracy and precision of HR measurements performed using the tested vPPG technology and a smartphone camera by comparing them with reference values obtained from a simultaneously recorded electrocardiogram (ECG).

## METHODS

### Subjects

We recruited 38 adult volunteers aged 20 to 43 years (median 25 years), of whom 20 were females. The subjects were generally healthy and in particular did not suffer from any cardiovascular disease. The exclusion criteria (which ultimately did not have to be applied) were as follows: arrythmia (other than sinus bradycardia or tachycardia), neurologic disorders in the form of spontaneous head tremor, inability to keep the head in the required position during measurement, respiratory disorders such as irregular or shallow breathing, facial shape deformation, or extensive damage, wound, burn, dressing or disease of the facial skin. The study was approved by the Bioethics Committee of the Wroclaw Medical University (approval number 227/2022), and written informed consents have been obtained from all study participants. Due to technical issues with some ECG signals (excessive noise or artefacts), we had to exclude data from three subjects, and hence the final analysis was based on data from 35 subjects (17 females). See Table 1 for the characteristics of the study participants.

**Table 1.**
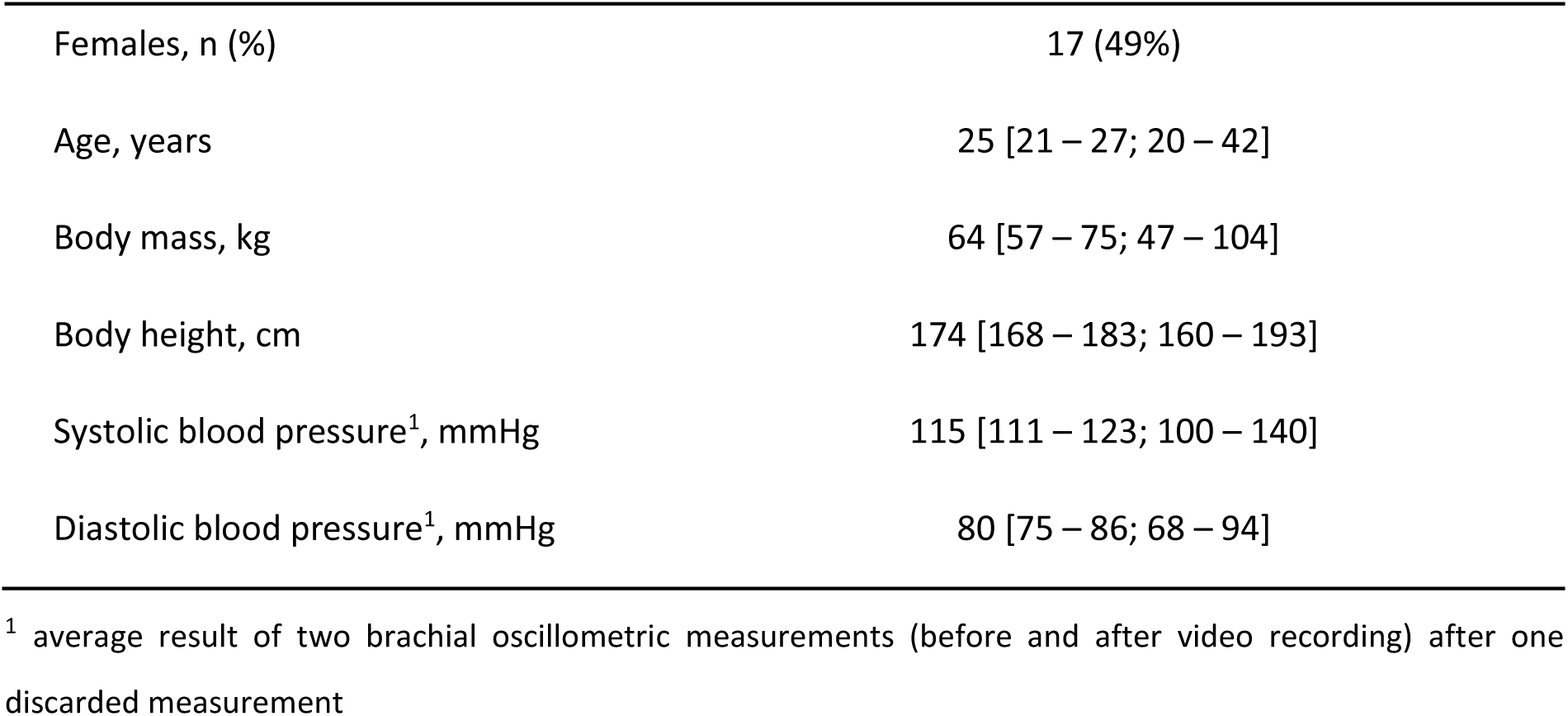
Characteristics of study participants (n = 35). Continuous variables presented as median [interquartile range; full range].

|  |  |
| --- | --- |
| Females, n (%) | 17 (49%) |
| Age, years | 25 [21 – 27; 20 – 42] |
| Body mass, kg | 64 [57 – 75; 47 – 104] |
| Body height, cm | 174 [168 – 183; 160 – 193] |
| Systolic blood pressure<sup>1</sup>, mmHg | 115 [111 – 123; 100 – 140] |
| Diastolic blood pressure<sup>1</sup>, mmHg | 80 [75 – 86; 68 – 94] |

<sup>1</sup> average result of two brachial oscillometric measurements (before and after video recording) after one discarded measurement

### Study protocol

The subjects were asked not to engage in any strenuous physical activity before participating in the study. During the study, they remained seated the entire time, with the back supported. To ensure that measurements were taken under resting conditions, video recordings were started approximately 5 min after all data acquisition devices were connected and set up. Arterial blood pressure was measured twice before the video recording (at a 1 min interval) and once immediately after the video recording. The subjects were asked to remain steady during the video recording, to refrain from speaking, and to breathe normally. Each participant was measured once in a resting (sitting) condition. This measurement was followed by two other measurements (in other conditions or with a specific breathing pattern) that are beyond the scope of this study.

### Tested parameters

We studied the following HR values provided by the tested technology based on a 1-min facial vPPG measurement:

1. HR averaged over 1 minute, i.e. the average HR from the entire measurement;
2. multiple HR values averaged over 10-sec periods – these short-term average HR values (49 values per measurement) are calculated and provided every 1 s during the measurement (starting from the 11^th^ second until the 59^th^ second) based on the preceding 10-sec period with a 1-sec delay (e.g. at time t = 11 s, the calculated value corresponds to the average HR in the period between t = 0 and t = 10 s);
3. multiple HR values averaged over 4-sec periods – these instantaneous HR values (54 values per measurement) are calculated and provided every 1 s during the measurement (starting from the 6^th^ second until the 59^th^ second) based on the preceding 4-sec period with a 1-sec delay (e.g. at time t = 6 s, the calculated value corresponds to the average HR in the period between t = 1 and t = 5 s);

### Equipment

Video recordings of participants’ faces were taken with the front camera of a mobile phone (Samsung Galaxy A22) mounted on a tripod at a distance of about 50 cm from the face and at a height adjusted to each participant (see Fig. 1). The camera parameters were as follows: f/2.2, sensor resolution 13 MP, video resolution 1080p, frame rate 30 fps. In parallel, a 1-lead chest ECG was recorded continuously using the Bio Amp ML132 module and the PowerLab data acquisition system (ADInstruments, Dunedin, New Zealand) with the following configuration of the three electrodes (Einthoven Lead I configuration): red (positive) on the right collarbone, white (negative) on the left collarbone, and black (reference) on the abdomen, to the left of the navel. Blood pressure was measured using a validated automatic upper arm blood pressure monitor (Omron M4 Intelli IT). Additionally, for other research purposes (not analysed in the present study), we monitored also blood pressure on the finger, arterial blood oxygen saturation (with an ear pulse oximeter), and breathing rate (using a chest belt detecting changes in chest circumference).

**Fig. 1.**
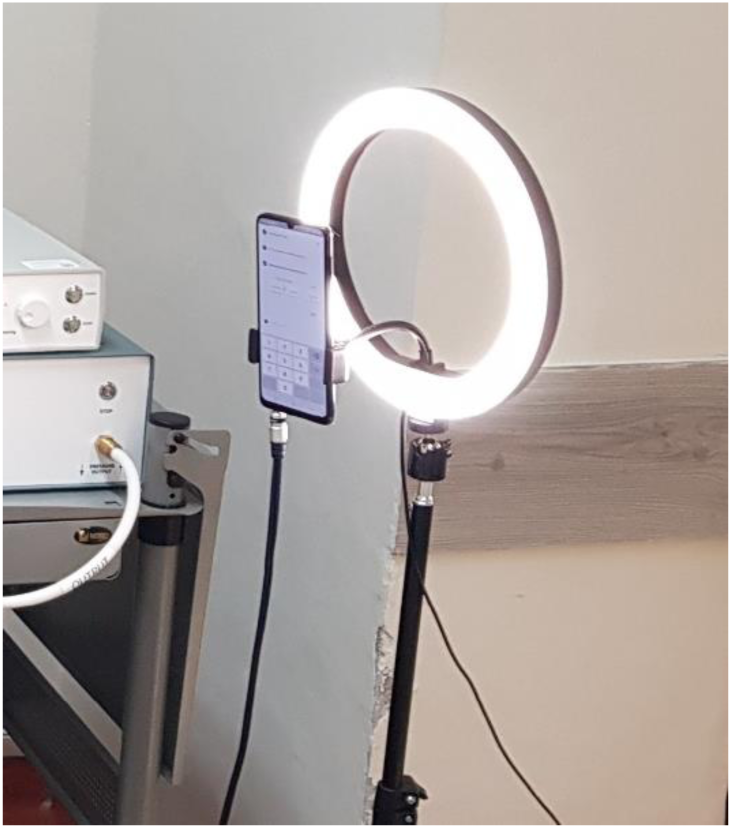
Measurement setup. The subject was seated approximately 50 cm from the phone with the back supported. The phone was mounted on a tripod at head level with an LED lamp in the background.

### Lighting

The measurements were taken during the day, but considering that they took place in a laboratory located in the semi-basement, with occasionally limited access to daylight (especially in cloudy conditions) and potential shadows cast by nearby trees and/or vehicles passing on a nearby street, to keep the light conditions constant, we decided to have the window blinds closed and the ceiling lights on during all measurements, regardless of the time of day or weather. Additionally, to ensure a sufficient level of light illuminating participants’ faces, we used a ring-shaped LED lamp (with the light colour corresponding to natural light) mounted on the tripod, with the phone in the centre and in front of the lamp (see Fig. 1).

### Video collection and processing

Video recordings were performed using a special mobile app developed and provided to us by MX Labs. This research app was designed to record a 2-min facial video that is then sent to the MX Labs cloud server to be processed and analysed by the Shen.AI Vitals algorithms to provide estimates of various physiological parameters, including average HR from selected time windows of the recorded video. In the publicly available app from MX Labs (called Heart Monitor), the recording/measurement is limited to 1 minute, and hence in the present study all analyses were done using only the first minute of the recorded video (thus ignoring the second minute). In the Heart Monitor app, all data processing and analysis takes place on the mobile device instead of on the cloud server, with the final results (including average HR) displayed to the user after the measurement and short-term HR values displayed during the measurement and updated every 1 second. In our study, which employed the research app, both participants and laboratory staff were blinded to the results of video measurements. These results (for all participants) were calculated on MX Labs cloud server using the Shen.AI Vitals algorithms and provided to us by MX Labs only after the study was completed in a digitalized tabular form. In this way, after calculating the reference HR values from the recorded ECG signals, we were able to compare HR values obtained independently by the two methods. In particular, we were able to compare the multitude of short-term HR values, which would have not been possible if we were using the normal (non-research) app, where these values are displayed every 1 second during the measurement and are not available after the measurement. Moreover, in this research app, a special audio signal (a beep) was sent by the app to the PowerLab system at the beginning and end of each video recording to facilitate synchronization of reference ECG signal with video measurement, which was crucial for our study.

### ECG processing

ECG signals recorded with the PowerLab data acquisition system (ADInstruments) were exported to LabChart 8 software (ADInstruments), which was used for automatic detection of QRS complexes and calculation of time intervals between successive heartbeats (R-R intervals). We used the LabChart default (human) settings for the QRS detection algorithm. In a few cases, where automatic detection of QRS complex failed, R peaks were marked manually in the LabChart interface. As mentioned earlier, in three subjects the ECG signals were very noisy and contained various artefacts, which prevented detection of most QRS complexes, and therefore data from these subjects were excluded from the analysis.

The calculated R-R intervals (in ms) along with their time stamps (the times of their ends) were exported from LabChart to a text file, which was subsequently processed as follows using a script in MATLAB (The Mathworks Inc., USA). First, we selected all R-R intervals that were entirely contained within the 60-sec period corresponding to the first minute of the video recording (based on the aforementioned audio signal that was also exported to the same text file). Second, we corrected occasional wrongly-identified R-R intervals (caused by falsely detected additional R-peaks that were merely local noise, as confirmed by visual inspection of the ECG signals) – this was done by combining two adjacent R-R intervals whenever their total duration was between 80% and 120% of the median duration of all R-R intervals identified within the 60-sec period of interest.

### Calculation of reference HR values

Average HR (in beats per minute, bpm) was calculated as the reciprocal of the mean duration of all R-R intervals in the analysed 60-sec period. Similarly, short-term (10-sec) or instantaneous (4-sec) HR values were calculated as the reciprocal of the mean duration of all R-R intervals in the given period, i.e. in the previous 10 or 4 seconds, respectively, with a 1.3-sec delay. For instance, the reference 4-sec HR average corresponding to the value provided by the tested technology at time t = 6 s was calculated as the reciprocal of the mean duration of all R-R intervals contained in the period between t = 0.7 s and t = 4.7 s. The additional 0.3-sec delay was applied to account for the typical time shift between the R-peaks in the ECG signal and the pulsation peaks in the facial vPPG signal, so that the R-R intervals used to calculate the reference short-term or instantaneous HR values matched the time intervals between the peaks in the vPPG signals considered by the tested technology. All reference HR values were rounded similarly as the tested values, i.e. to the nearest whole number.

### Statistics

HR values obtained using the tested technology were compared with the reference ECG-based values by calculating mean errors (ME), standard deviations of errors (SD), mean absolute errors (MAE), and root-mean-square errors (RMSE) in absolute terms as well as in relative terms, i.e. with respect to the reference values. Additionally, for all analysed HR parameters, the correlation between the tested and reference values was assessed using the Pearson correlation coefficient (r) and the agreement between them was visualized using scatter plots. Statistical significance was set at *P* = 0.05.

## RESULTS

According to ECG, the median value of the average HR among the study participants was 75 bpm (interquartile range 72–81 bpm, total range 52–98 bpm). In Table 2 we show various measures of accuracy and precision of HR values estimated by the tested vPPG technology (compared to ECG-based values), in both absolute and relative terms, i.e. with respect to reference values. In particular, we assessed the average HR from the entire measurement (60 s) and multiple HR values averaged over shorter periods, i.e. 10 s (short-term HR) or 4 s (instantaneous HR). Regardless of the HR averaging time, the mean errors were close to 0.1 bpm. The mean absolute errors (MAE) were approximately 0.1, 0.2, and 0.4 bpm for HR averaged over 60-sec, 10-sec, and 4-sec periods, respectively. For the average HR from the entire measurement, there were no errors larger than 1 bpm (see Table 3). For short-term or instantaneous HR values, i.e. average values from 10-sec or 4-sec periods, the errors did not exceed 1 bpm in 99.8% and 94.5% of cases, respectively. The maximal errors amounted to 3 bpm for 10-sec averaging periods (1 case out of 1715) and 4 bpm for 4-sec periods (4 cases out of 1890).

**Table 2.**
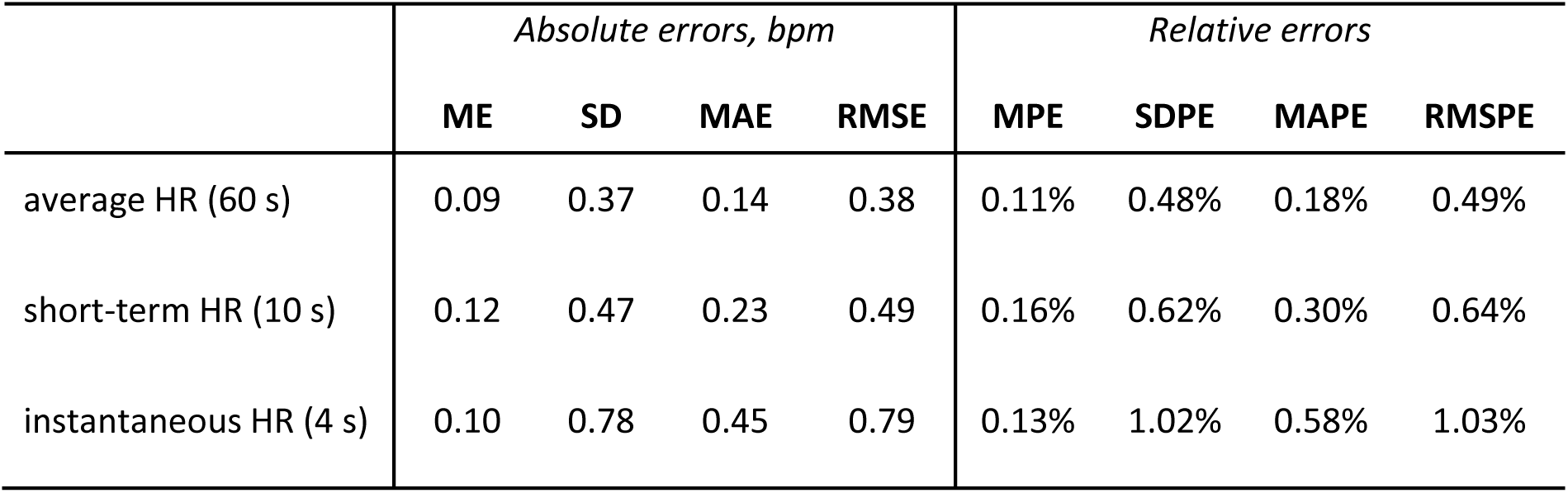
Measures of accuracy and precision of heart rate (HR) estimated by the Shen. AI Vitals technology as compared with ECG-based values, expressed in absolute terms (in beats per minute, bpm) or in relative terms (in percentage of ECG-based values). Abbreviations: ME – mean error, SD – standard deviation of errors, MAE – mean absolute error, RMSE – root-mean-square error, MPE – mean percentage error, SDPE – standard deviation of percentage errors, MAPE – mean absolute percentage error, RMSPE – root-mean-square percentage error.

**Table 3.** Distribution of absolute errors in heart rate (HR) values estimated by the Shen. AI Vitals technology as compared with reference values obtain from simultaneously recorded electrocardiogram. The data shows errors in HR averaged over 60 s as well as over shorter periods (10 s or 4 s).

| Error | HR (60 s) | HR (10 s) | HR (4 s) |
| --- | --- | --- | --- |
| 0 bpm | 30 (85.7%) | 1322 (77.1%) | 1181 (62.5%) |
| 1 bpm | 5 (14.3%) | 389 (22.7%) | 605 (32.0%) |
| 2 bpm |  | 3 (0.2%) | 79 (4.2%) |
| 3 bpm |  | 1 (0.1%) | 21 (1.1%) |
| 4 bpm |  |  | 4 (0.2%) |
| $\leq 1$ bpm | 35 (100%) | 1711 (99.8%) | 1786 (94.5%) |
| $\leq 2$ bpm | | 1714 (99.9%) | 1865 (98.7%) |
| $\leq 3$ bpm | | 1715 (100%) | 1886 (99.8%) |
| $\leq 4$ bpm | | | 1890 (100%) |

Fig. 2 presents the HR values obtained using the tested technology plotted against the reference values from ECG. The correlation between HR values from the two methods was very strong as given by the Pearson correlation coefficients: r = 0.992 for 60-sec HR, r = 0.999 for 10-sec HR, and r = 0.997 for 4-sec HR (*P* < 0.001 in all cases). For individual subjects, MAE varied between 0.2 and 1.0 bpm for 4-sec HR, and between 0.1 and 0.5 bpm for 10-sec HR (see Fig. 3), and the two MAEs were strongly correlated with each other (r = 0.817, P < 0.001).

**Fig. 2.**
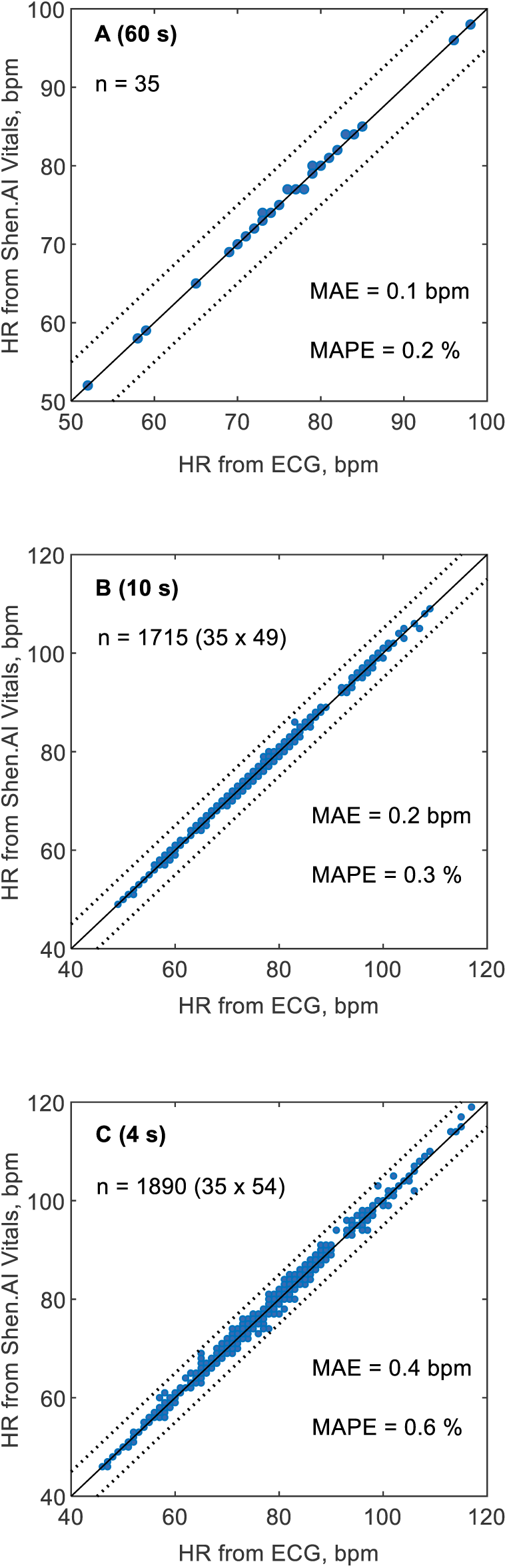
Heart rate (HR) values estimated in 35 subjects using the Shen.AI Vitals facial vPPG technology plotted against reference values obtained from simultaneously recorded electrocardiogram (ECG). **A)** average HR values from 60-second measurements, **B)** short-term HR values averaged over 10-second periods (49 values per subject), **C)** instantaneous HR values averaged over 4-second periods (54 values per subject). Dotted lines indicate error limits of ± 5 beats per minute (bpm). Abbreviations: MAE – mean absolute error, MAPE – mean absolute percentage error.

**Fig. 3.**
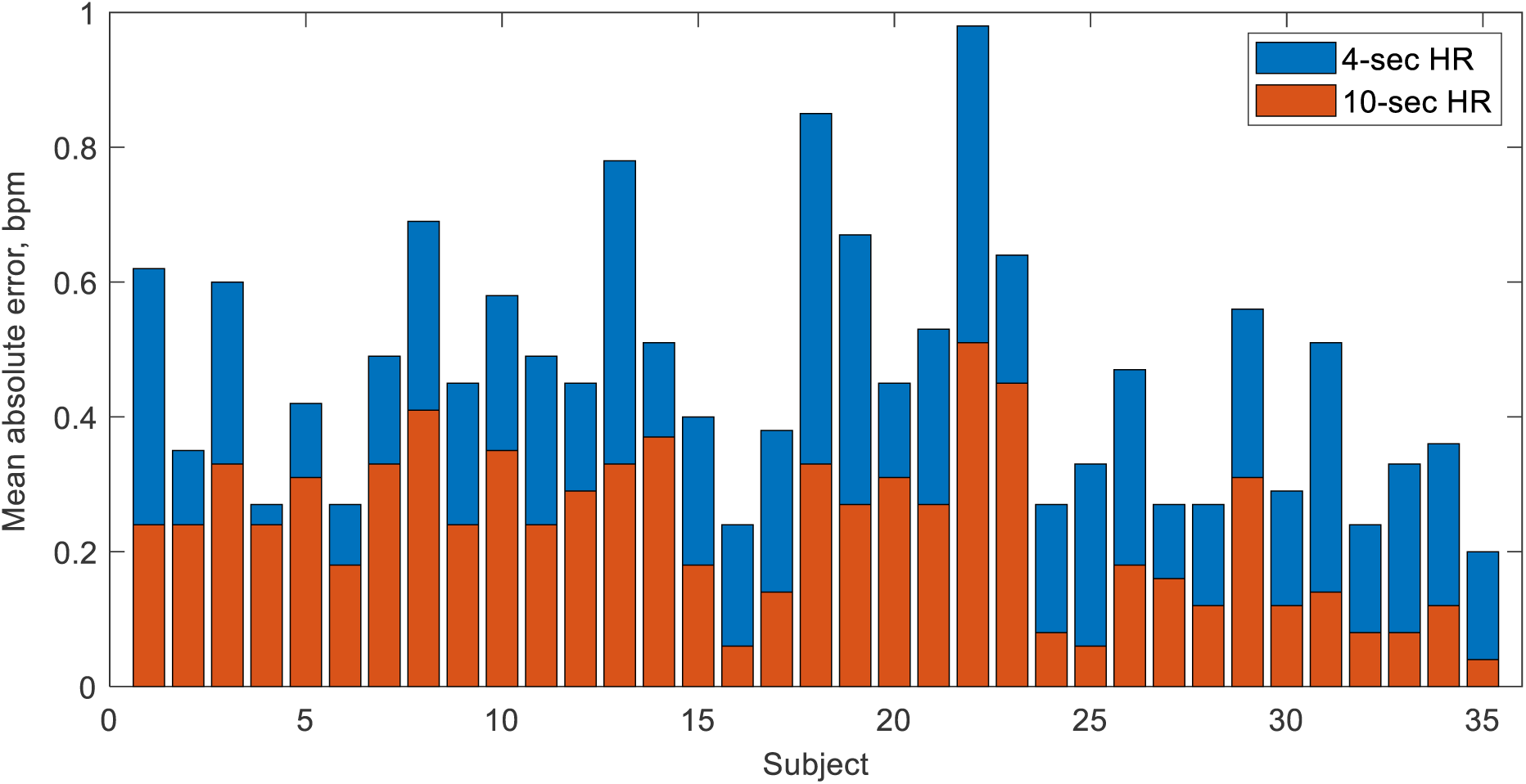
Mean absolute errors (per subject) in heart rate (HR) values estimated by the Shen.AI Vitals technology in multiple 4-sec periods (blue bars, top) and 10-sec periods (red bars, bottom) during 60-sec video recording of the face, as compared to electrocardiogram-based values. For each subject, the two mean absolute errors correspond to the means of 54 and 49 values for HR averaged over 4-sec and 10-sec periods, respectively (see Methods for details). All bars start at 0.

As shown in Fig. 4 (right panel), the instantaneous HR values calculated from ECG (averaged over 4-sec periods) showed different levels of variation during the 60-sec measurements in individual subjects (e.g. related to breathing patterns). In most subjects, those instantaneous HR values varied within approximately ±5 bpm, with one subject showing particularly high HR variation (between 80 and 117 bpm). Across all subjects, we observed instantaneous HR values between 46 and 117 bpm. For HR values averaged over 10-sec periods, as expected, the variation was accordingly lower, with low-frequency trends visible in some subjects (see left panel in Fig. 4).

**Fig. 4.**
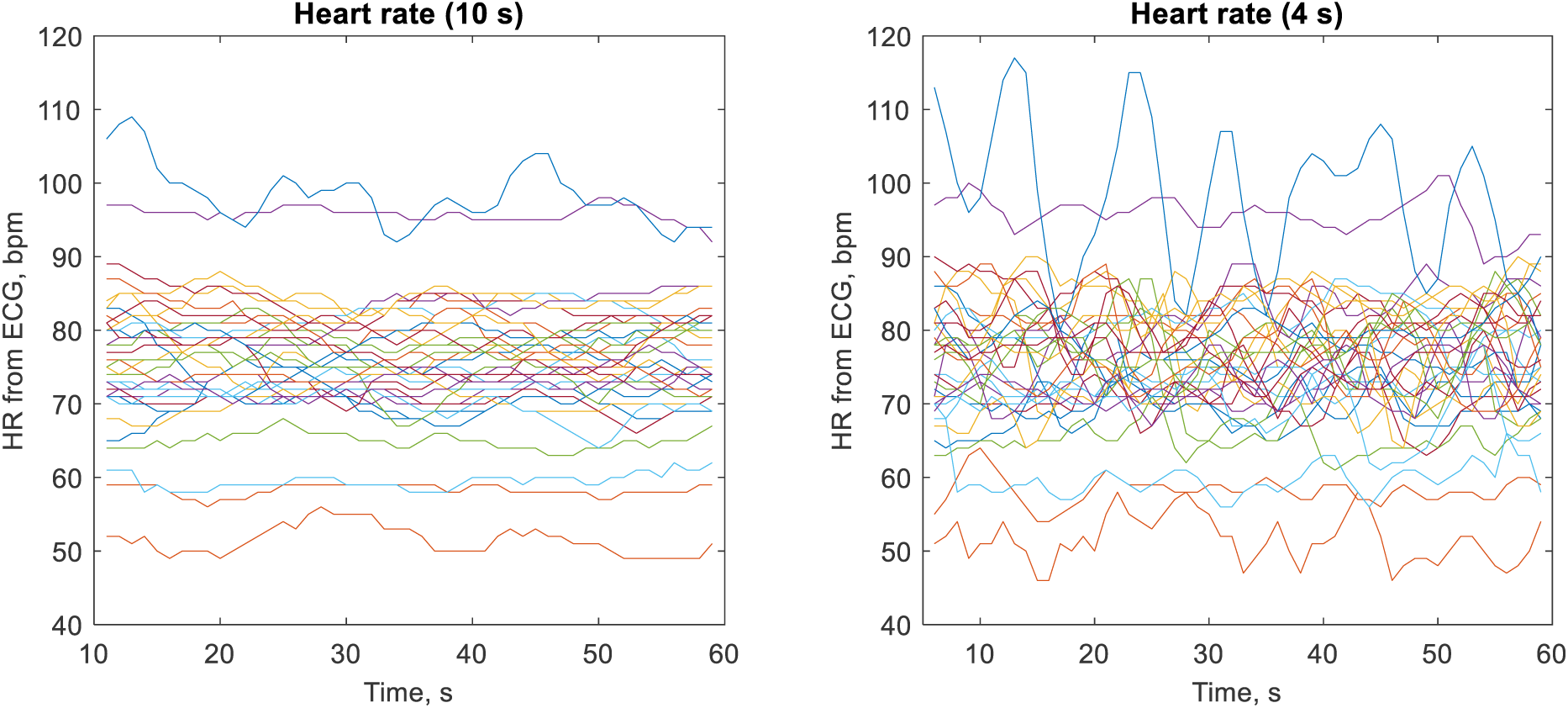
Heart rate (HR) values obtained from electrocardiogram (ECG) averaged over 10-sec periods (left panel) and 4-sec periods (right panel) during 60-sec measurements. Each line represents one of the 35 studied subjects (with the same colour used in both panels), with HR values updated every 1 s (see Methods for details).

## DISCUSSION

In this study on 35 young, healthy volunteers, we observed a very high agreement between HR values estimated by the tested vPPG technology and the ECG-based values, particularly for the average HR from the entire 60-sec measurement, for which in all subjects the errors did not exceed 1 bpm and in 86% of them there was virtually no difference between the vPPG-based and ECG-based average HR. The mean absolute error (MAE) of these measurements was only around 0.1 bpm, whereas the root-mean-square error (RMSE) was around 0.4 bpm. Even for HR averaged over much shorter periods, i.e. multiple HR values averaged over 10-sec or 4-sec periods, the agreement was also very high, with errors not exceeding 1 bpm in 99.8% and 94.5% of cases, respectively, and the RMSE of 0.5 bpm and 0.8 bpm, respectively.

### Standards

According to the standard ANSI/AAMI EC13:2002 (R2007) “Cardiac Monitors, Heart Rate Meters, and Alarms” [29], later superseded by ANSI/AAMI/IEC 60601-2-27:2011 (R2016) [30], the HR measurement error should not be greater than ±10% or ±5 bpm, whichever is greater. It should be noted, however, that this standard applies to ECG-based devices and it specifically excludes PPG-based devices. Nevertheless, given the lack of more appropriate standards, it is frequently used as a reference when assessing the accuracy of PPG or vPPG-based HR monitors [31, 32]. In our study, we observed no errors in HR greater than 4 bpm or 10%, even for HR averaged over 4-sec periods, despite the relatively high variation in these instantaneous HR values during 60-sec measurements. We investigated 1890 of such instantaneous HR values (54 in each of the 35 studied subjects) with a relatively wide range of values (between 46 and 117 bpm, according to ECG), and we observed errors of 4 bpm only in 4 cases (0.2%). These 4-sec average HR values can be in fact treated as instantaneous HR in line with the expert statement of the INTERLIVE Network regarding the validity of consumer wearable HR monitors, which indicates that instantaneous HR values should be averaged in time windows not longer than 5 s [33]. The high level of accuracy of these instantaneous HR values observed in our study suggests that vPPG technology has the potential to be used in biofeedback applications [34–37].

In a less strict standard, ANSI/CTA-2065 “Physical Activity Monitoring for Heart Rate and Related Measures” [38], developed by the Consumer Technology Association (CTA) for wearable devices for continuous HR measurements during physical activity, the accuracy criterion is that the mean absolute percentage error (MAPE) should not be greater than 10% for all measurements during the protocol proposed in that standard (pooled for all participants). Although we did not study a device for continuous HR measurements during physical activity, the short-term HR (averaged over 10 s) and the instantaneous HR (averaged over 4 s) are provided by the tested technology every 1 s, and hence they can be treated as a kind of continuous HR measurements, which could potentially last longer than the current measurement time (60 s). We observed MAPE of only around 0.3% and 0.6% for HR averaged over 10-sec and 4-sec periods, respectively, thus markedly lower from the level required by the above standard, although we emphasize that we studied sedentary and inactive subjects, which of course is of great importance here.

### Time shift between ECG and vPPG peaks

Note that for HR values averaged over 10-sec and 4-sec periods, the reference ECG-based values were averaged over the corresponding periods lagged by 0.3 s (see Methods) to account for the fact that the peaks in the vPPG signals are delayed with respect to R peaks in ECG used for heartbeat detection. For simplicity, in all subjects we used the same time shift of 0.3 s, which was approximately the typical time shift observed in our dataset. However, this time shift differed to some extent both between and within the subjects (over the course of measurement). This means that in some cases, the reference ECG-based HR values could have been based on a slightly different number of time intervals, e.g. the number of R-R intervals in the given 4-sec period could have been lower or higher by one compared to the number of peak-to-peak time intervals in the vPPG signal analysed by the tested technology. We believe that this may explain part of the differences between the HR values estimated by the tested technology and the reference values, which could be avoided or at least mitigated by using personalized time shifts instead of the fixed value of 0.3 s. Therefore, we expect that the accuracy of those short-term or instantaneous HR values provided by the tested technology could be even higher. For HR averaged over the entire measurement (60 s), we did not use any time shift when calculating the reference values from ECG, as in this case such a time shift has a negligible impact. Again, accounting for such a time shift could lead to even better results in terms of accuracy of vPPG-based average HR values.

### Previous studies

As far as we know, the most data on the accuracy of vPPG-based measurements of HR are available for the Lifelight technology. In their largest study (VISION-D) involving over 10,000 HR measurements in more than 5,700 subjects, the mean error (ME) was 0.3 bpm with the standard deviation of errors (SD) of 4.0 bpm [39], thus markedly higher than in our study (0.1 and 0.4 bpm, respectively), although their study was not only much larger than ours but included a very diverse population of patients and healthy volunteers, with a wide range of age, health condition, and skin tone, and a wider range of HR (32–183 bpm). Note also that they used reference HR values from a sphygmomanometric measurement performed during 1-min vPPG measurement, which on the one hand makes the comparison less reliable than when comparing with ECG-based reference values, but on the other hand could explain higher errors. In a more recent, smaller study on the Lifelight technology, with 57 measurements in 19 participants and ECG-based reference HR values, the ME was higher in absolute terms (-0.6 bpm), but the SD was lower (1.8 bpm) [40], and hence those measurements were more precise than in their previously mentioned large study. In another study that investigated 1-min vPPG measurements of HR in 45 seated subjects (with relatively low variation in terms of age and skin tone), MAE was reported at 1.7 bpm, with SD of 2.1 bpm [41]. In a study on 9 young, healthy volunteers with 8 measurements per subject, the results were only slightly worse than in our study, with ME of -0.2 bpm and SD of 0.6 bpm, as compared to ECG-based values, although that study involved 2-min measurements with various breathing rates and depths [42].

As far as shorter measurements are concerned, Hassan et al. compared several vPPG methods for 30-sec measurements of HR in 45 healthy volunteers and obtained relatively high errors, with ME ranging from 3.5 to 11.0 bpm and SD ranging from 3.5 to 8.2 bpm, although they studied measurements in a more natural environment, with uneven illumination of the participants’ faces and possible shadows or reflections, with a distance of 0.8 m between the camera and the face [43]. In a study on 25 subjects with 20-sec vPPG measurements using a smartphone camera with flash, Sanyal and Nundy reported ME of -0.1 bpm and SD of 4.2 bpm [44]. In a study looking at 10-sec HR measurements (during 2-min video recordings) in 22 subjects, ME was -0.2 bpm and SD was 1.8 bpm [45]. Tran et al. investigated HR averaged over 8-sec periods during 1-min video recordings in 10 subjects and found ME of -0.5 bpm and RMSE of 5.7 bpm, although their dataset included measurements taken at various distances between the camera and the face, various lighting conditions, and intentional head movements [46]. For an overview of accuracy of vPPG-based measurements of HR, including older studies with low-resolution and/or low-framerate cameras, see the reviews by Rouast et al. [47] or Molinaro et al. [19].

### Limitations

Our study had certain limitations. First, we studied a relatively small and homogeneous sample, which included only white subjects, and hence our results are not necessarily generalizable to dark-skinned individuals, in whom PPG signals have usually lower quality due to larger amount of light being absorbed by the skin rather than reflected [48, 49]. Second, we studied subjects in a sitting position who, in accordance with our request and requirements of the tested technology, kept their heads relatively still during the measurements and refrained from speaking. Although the tested technology includes a face tracking algorithm to account for possible head movements or changes in facial expressions, we have not investigated how these factors may affect measurement accuracy, which would require a separate study with intentional head movements and/or changes in facial expressions.

Finally, our study was conducted in a laboratory setting, with controlled lighting conditions and the smartphone camera positioned at the participant’s head level. Although these conditions were similar to those typically seen in studies of this type, which facilitates comparison of results across study participants, our results may not necessarily generalize to other, more natural settings, with potentially lower facial illumination levels (on the other hand, higher illumination levels could potentially lead to better results).

## CONCLUSIONS

In a group of predominantly young, white participants in a resting (sitting) position, the tested facial vPPG technology provided highly accurate heart rate measurements, both in terms of the average HR from the entire 60-sec measurement as well as short-term HR values averaged over 10-sec periods and instantaneous HR averaged over 4-sec periods, with errors not exceeding 1 bpm in 100.0%, 99.8%, and 94.5% of cases, respectively. Our results confirm the feasibility of using a smartphone camera and facial vPPG technology to measure resting HR, although this should be further confirmed in a study involving a larger group of participants with greater diversity in age, skin tone and lighting conditions.

## Data Availability

Anonymized heart rate data analyzed in the present study are available upon reasonable request to the corresponding author.

## Acknowledgements

MX Labs has provided us with a special (research) version of its mobile application as well as the smartphone and the tripod with an LED lamp used in the study.

## Conflicts of interest

The study was commissioned by MX Labs and conducted under a contractual agreement between MX Labs and Wroclaw Medical University. LP and TO report personal fees from MX Labs. Other authors have no conflicts of interest.

## Author contributions

L. Pstras: Conceptualization, Methodology, Software, Data curation, Formal Analysis, Writing – original draft, Writing – review & editing, Visualization.

T. Okupnik: Investigation, Data curation.

B. Ponikowska: Resources, Supervision.

B. Paleczny: Conceptualization, Supervision, Writing – review & editing.

## Notes

### Funding Statement

Wroclaw Medical University received a payment from MX Labs to support the present study.
LP and TO report personal fees from MX Labs.

### Author Declarations

The Bioethics Committee of the Wroclaw Medical University gave ethical approval for this study (approval number 227/2022).

